# Mathematical models of COVID-19 vaccination in high-income countries: A systematic review

**DOI:** 10.1101/2024.03.24.24304676

**Authors:** Eleanor Burch, Saher Aijaz Khan, Jack Stone, Asra Asgharzadeh, Joshua Dawe, Zoe Ward, Ellen Brooks-Pollock, Hannah Christensen

## Abstract

**Objectives:** Since COVID-19 first emerged in 2019, mathematical models have been developed to predict transmission and provide insight into disease control strategies. A key research need now is for models to inform long-term vaccination policy. We aimed to review the variety of existing modelling methods, in order to identify gaps in the literature and highlight areas for future model development.

**Study design:** This study was a systematic review.

**Methods:** We searched PubMed, Embase and Scopus from 1 January 2019 to 6 February 2023 for peer-reviewed, English-language articles describing age-structured, dynamic, mathematical models of COVID-19 transmission and vaccination in high-income countries that include waning immunity or reinfection. We extracted details of the structure, features and approach of each model and combined them in a narrative synthesis.

**Results:** Of the 1109 articles screened, 47 were included. Most studies used deterministic, compartmental models set in Europe or North America that simulated a time horizon of 3.5 years or less. Common outcomes included cases, hospital utilisation and deaths. Only nine models included long COVID, costs, life-years or quality of life-related measures. Two studies explored the potential impact of new variants beyond Omicron.

**Conclusions:** This review demonstrates a need for long-term models that focus on outcome measures such as quality-adjusted life years, the population-level effects of long COVID and the cost-effectiveness of future policies – all of which are essential considerations in the planning of long-term vaccination strategies.

## Introduction

The rapid development and distribution of multiple COVID-19 vaccines has allowed non-pharmaceutical interventions to be largely removed worldwide while controlling the number of COVID-19 hospitalisations and deaths. Although COVID-19 hospitalisation and death rates have decreased considerably since the peak of the pandemic, COVID-19 remains an ongoing public health challenge, and there is a continued need to monitor infection levels and update long-term vaccination programmes. Vaccines provide imperfect protection, despite being the primary public health strategy for reducing COVID-19-related morbidity and mortality,^1^ and immunity from both vaccination and previous infection wanes over time.^2^ Furthermore, immune escape can occur in the case of new variants, as has been shown for Omicron, which affected vaccine effectiveness further.^3^

Mathematical modelling is an important tool to predict the transmission of SARS-CoV-2 and inform the development of vaccination programmes.^4^ Different types of models are used to explore the impacts of vaccinating population groups and scheduling multiple doses.^5,6^ Models vary in terms of their structure, features and assumptions, with the public health question and study aims determining which model design should be used. For example, susceptibility to severe COVID-19 disease and contact rates vary by age, and in order to compare long-term vaccination strategies targeted at specific risk groups, it is important to include age structure in models. Mathematical modelling of SARS-CoV-2 has evolved over time in response to emerging evidence. Early SARS-CoV-2 mathematical models generally did not include immune decay.^7–9^ This has been increasingly captured in models, although the exact dynamics of waning immunity are still uncertain, and therefore incorporated into models using different approaches.

Systematic reviews of SARS-CoV-2 mathematical modelling studies have mostly concerned models published in 2020 and have focused on model results and predictions, for example to estimate key epidemiological parameters.^10–15^ The use of modelling to inform policy around disease control strategies in healthcare settings has been demonstrated,^13^ and a synthesis of model results was used to recommend vaccine prioritisation strategies globally.^15^ A review of models set in Sweden studied the validity and accuracy of model predictions compared to actual outcomes.^14^ Reviews that have considered modelling methods have primarily focused on basic details such as the model type, compartments, setting, outcomes and input parameters, as well as some epidemiological features in the models including the importance of asymptomatic transmission.^10,11,13,15,1612^ To date, no systematic review has investigated approaches to modelling SARS-CoV-2 vaccination and waning immunity, that includes recent models exploring long-term control strategies and booster vaccine rollout.

Here, we present results from a systematic review of age-structured mathematical models of SARS-CoV-2 transmission set in high-income countries that include vaccination and waning immunity. The aim of this study was to compare and provide a narrative synthesis of the various modelling methods, and to identify where future model development is required to inform long-term vaccination strategies.

## Methods

### Search and selection

This systematic review was conducted and reported following the Preferred Reporting Items for Systematic Reviews and Meta-Analyses (PRISMA) guidelines (supplementary file 4). The study protocol was registered on Prospero on 17 August 2022 (registration ID: CRD42022353757, available from: https://www.crd.york.ac.uk/prospero/display_record.php?ID=CRD42022353757). The protocol has since been amended to add details of review team members, update the end date of the review, and update the search strategy and eligibility criteria for the reasons explained in the protocol. We carried out a search of PubMed, Embase and Scopus on 6 February 2023. The search contained a combination of MeSH/Emtree terms and key words within article titles including “COVID-19”, “SARS-CoV-2”, “vaccination” and “model”. The full search strategy is available in supplementary file 1. We restricted the search to papers that were published in English, in peer-reviewed journals, between 1 January 2019 and 6 February 2023, and excluded animal studies.

We included studies describing age-structured, dynamic, mathematical models of COVID-19 in human populations, that incorporated vaccination and any form of waning immunity or reinfection (either in the standard scenario or within sensitivity analyses). We defined this as the waning of either natural or vaccine-induced protection over time, or the possibility of reinfection of previously infected individuals which was not solely due to immune escape of new variants. Definitions of key modelling terms are shown in table 1. Eligible studies were set in general population settings in high-income countries, according to the World Bank 2023 classification.^18^ The limitation to high-income settings was agreed to make the review manageable, as the data extraction of modelling methods would not have been practical for the full extent of published models globally. Studies describing only animal, statistical or within-host models were excluded, as well as mathematical models that did not involve dynamic transmission or age stratification. We also excluded reviews, studies that described purely theoretical models with no evaluation of epidemiological or policy implications, and studies where a real-world setting was not specified.

**Table 1.** Definitions of key terms used in infectious disease modelling.

| Term | Definition |
| --- | --- |
| Dynamic transmission model | An infectious disease model that simulates transmission in a population over time. |
| Compartmental model | The modelled population is divided into compartments representing the stages of an infectious disease. Individuals move between compartments as they pass through stages. Typical models include compartments for Susceptible, Infected and Recovered. |
| Agent-based model | Individuals are modelled as agents that interact with each other according to a set of rules.<sup>17</sup> Agents are assigned characteristics and behaviours, and can move between different states of infection. |
| Stochastic vs. deterministic models | Stochastic models use random variables to account for the element of chance in the spread of a disease. Deterministic models are fixed: given the same set of input parameters, the model output will always be the same. |
| Predictive vs. retrospective models | Predictive models are used to project the spread of disease in the future and evaluate the impact of disease control strategies. Retrospective models use historical data to simulate past events, sometimes including counterfactual analyses. |
| Immune escape | In this review we defined immune escape as pre-existing immunity from infection by one virus strain providing less protection against another, or vaccination granting varying levels of protection for different strains. |
| Time horizon | The duration of time over which a model is simulated. |
| “All-or-nothing” response | A response to vaccination wherein an individual develops either full or no immunity to infection depending on the vaccine effectiveness. This is in contrast to a “leaky” vaccination response in which all vaccinated individuals are equally less likely to become infected, depending on the vaccine effectiveness. |

The database search and de-duplication process using Microsoft Excel was carried out by a single reviewer [EB]. All screening of titles and abstracts was completed by EB using Rayyan^19^ and excluded articles were then crosschecked by SAK. Four authors [EB, SAK, JS, JD] contributed to the full-text screening, and each article underwent screening by two independent reviewers. Any disagreements were settled through discussion between the two reviewers. Once the screening process was complete, we searched the reference lists of included articles for other relevant papers. A search was also carried out using ResearchGate for eligible published papers describing the models used in the European COVID-19 Scenario Hub. We chose to include all studies describing the same model since changes were often made to the methods or model details.

### Data collection, synthesis and quality assessment

The data extraction and quality assessment procedures were completed in parallel by two independent reviewers [all authors contributed to this] and both sets were then compared for consistency. Once again, discrepancies were resolved through discussion. Missing data was requested from the study authors by email, and recorded as “not reported” when not supplied following contact. Where the same model was used in multiple papers we have included all versions of the model in this review.

Extracted data was recorded in an Excel spreadsheet and consisted of: study information, model type, structure and compartments, whether the model is deterministic or stochastic, whether the model is predictive or retrospective, model setting, age groups used, population stratification for vaccination, structure of the contact matrix, elements of COVID-19 biology and vaccination included in the model, time horizon of the study and inclusion of an economic evaluation. Supplementary file 1 contains details of any assumptions made and of the specific data extracted. Supplementary file 3 contains the template data extraction table. All results were pulled together in a narrative synthesis and model features were summarised.

To assess study quality, we modified the risk of bias tool developed by Harris et al. (details in supplementary file 1).^20^ We awarded each article a score between zero and two for each criterion, according to its suitability and clarity. We considered the model aims, structure, methods, interventions, assumptions, parameters, population and setting, and the fitting or validation methods used, as well as whether quality of data was considered and any uncertainty taken into account. In addition, we assessed whether the results were presented and discussed appropriately, and whether the funding sources and conflicts of interest were reported. Each paper had a total score out of 28 and was rated either “low” (score of 0-13), “medium” (14-18), “high” (19-22) or “very high” (22-28) accordingly. In order to combine the assessments made by the two independent reviewers, we compared the overall ratings and there was only considered to be a discrepancy if these did not match. In this case, an overall rating was agreed in a conversation between the two reviewers.

## Results

### Study selection

The searches identified 1641 studies, 1054 of which were distinct (figure 1). Title and abstract screening excluded 696 articles. Following full text screening of 358 studies, 40 articles were included, having met our eligibility criteria.^21–60^ A further six eligible studies were identified through screening reference lists of included studies,^61–66^ and one additional study was discovered through searching for papers describing the European COVID-19 Scenario Hub models,^67^ giving a total of 47 studies included in our review. Out of the 47 included studies, there were 40 unique models; five models were used in more than one paper.^31,35,38,40,42,46,47,50,55,59,61,63^ Key characteristics of the included models are shown in table 2 and the complete data extraction table is available in supplementary file 2.

**Figure 1.**
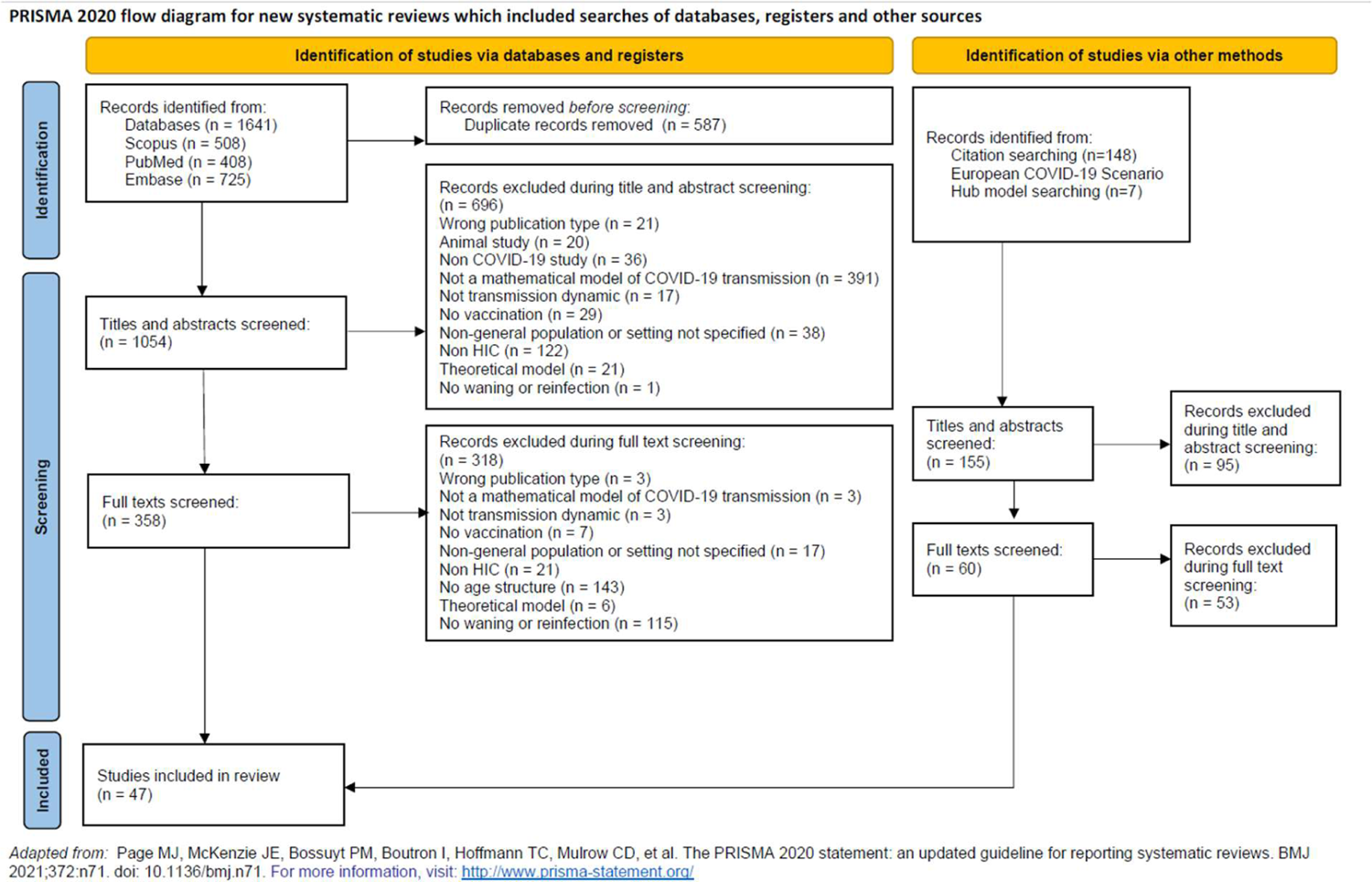
PRISMA flow diagram

**Table 2.**
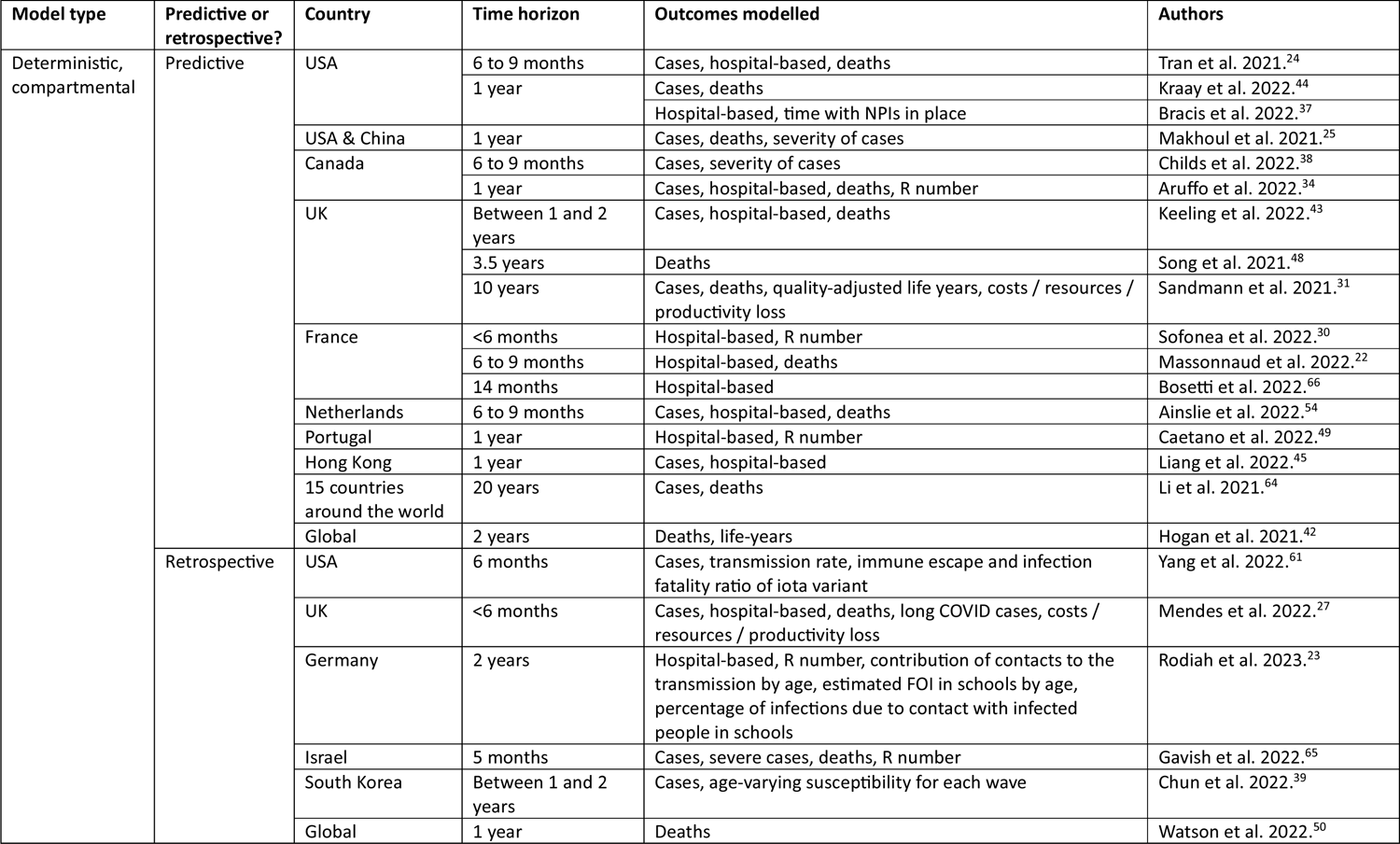
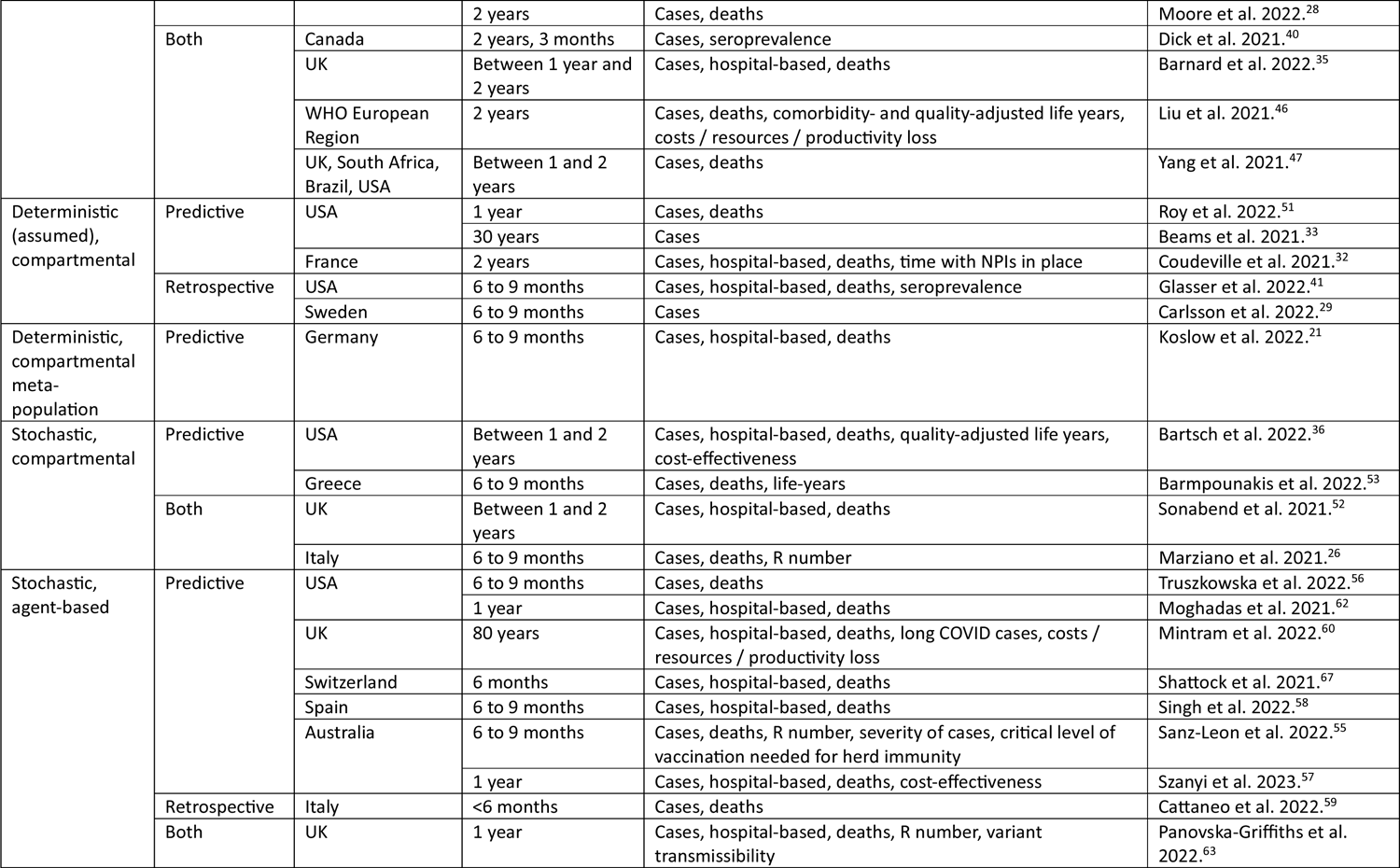
Key model characteristics for each study.

### Quality assessment

Overall, the studies scored highly in the quality assessment (supplementary file 2). Twenty papers were considered to be of “very high” quality, 20 “high” quality and seven “medium” quality. The total scores awarded by either of the independent reviewers ranged from 14 to 28. Studies were most frequently marked down for poor description or suitability of model validation and fitting methods, and for lack of explanation of the model structure and time horizon.

### Model type, time horizon and setting

Deterministic, compartmental models were used in the majority of studies (34/47), including one compartmental metapopulation model. Nine studies used stochastic, agent-based models, and four used stochastic, compartmental models. Most analyses (30/47) were predictive, ten were retrospective and seven both predictive and retrospective. We found that almost all studies (43/47) had a relatively short time horizon of up to 3.5 years, with an overall range of three months to 80 years. More than three quarters of the studies were set solely in North America or Europe (36/47), 19 of which included the UK or the USA, and three studies captured a global population. Most studies (38/47) described models at a country scale (i.e. the modelled population represented a whole country), whereas 10 were at a city or regional scale (including one study that did both^47^).

### Model outcomes

Most studies considered number of cases (including incidence, cumulative cases, and cases stratified by age, severity, strain, or separated into symptomatic/asymptomatic), hospital-based outcomes (including incremental, peak and cumulative admissions to hospital or ICU, bed-days and hospital system pressure) and deaths. Seven studies accounted for the effects of long COVID, and six of these also considered costs (either direct or indirect), resource use or cost-effectiveness of interventions. Five models incorporated either life-years, quality adjusted- or comorbidity adjusted-life years. Other outcomes modelled include the reproduction number, amount of time with non-pharmaceutical interventions (NPIs) in place and seroprevalence.

### Variants

From March 2022 onwards, studies began to specifically model the Omicron variant (10/47). Of the variants of concern that have emerged, the most common to be explicitly modelled in the reviewed papers was Delta, followed by Alpha. Some studies (7/47) used a single, generic set of parameters to describe the transmission and severity of SARS-CoV-2, without naming a specific strain. Only two studies explored the potential appearance of novel variants after Omicron. Immune escape was incorporated into just under half of the studies (23/47).

### Duration of immunity

The duration of immunity (both natural and vaccine-induced) ranged widely, from 90 days to lifelong. Some studies considered multiple durations of vaccine-induced immunity (16/47) or natural immunity (13/47) to account for uncertainty. A complete loss of protection over a period of time, where individuals returned to being fully susceptible, was included in some studies. Others described a partial loss of vaccine immunity within the simulation period – sometimes providing the half-life and sometimes a percentage reduction. Including sensitivity analyses, 14 studies did not consider waning of either one of: natural immunity, or vaccine-induced immunity. Moreover, recovered individuals were completely immune against all reinfection in eight models that had lifelong natural immunity. Another five models included reinfection but no dynamic waning, where recovered individuals had a chance of developing either full or no immunity against reinfection post-infection (all-or-nothing response).

## Discussion

Our systematic review of 47 modelling studies of COVID-19 vaccination in high-income countries demonstrated a shortage of long-term models (with a time horizon of more than 3.5 years) that consider long COVID, costs, or outcome measures that include quality of life. We also identified a lack of models that investigate the possible impact of novel variants of concern, which may have increased levels of transmissibility, immune escape or likelihood of severe disease. This type of model could provide useful insight for future preparedness. More than three quarters of included studies were set in North America or Europe.

There is increasing evidence that COVID-19 immunity decreases over time.^68^ Furthermore, protection from past infection against reinfection is imperfect, particularly against the Omicron variant.^69^ Of the 14 studies that did not incorporate waning of either natural immunity or vaccine-induced immunity, seven simulated a time period of one year or more, and eight did not include any reinfection of recovered individuals. These assumptions reduce the reliability of these models and their usefulness in the future.

Model requirements and key outcome measures have changed over time. In the early stages of the COVID-19 pandemic, when quick decisions needed to be made on short-term resource planning and vaccine distribution, these decisions were based on reducing hospitalisations and deaths, in order to minimise the impact on health services. A crucial requirement now is for models that can be used to inform long-term policy decisions, which therefore need longer time horizons and must take into account the cost-effectiveness of interventions or consider measures of disease burden that include both the quality and quantity of life (e.g. quality-adjusted life years (QALYs)). Such measures place more weight on the lives of young people and are an essential consideration in the planning of long-term vaccination programmes. For example, in the UK, the Joint Committee on Vaccination and Immunisation (JCVI) bases cost-effectiveness analyses on cost per QALY gained.

Long COVID is a growing public health issue and has a large impact on the day-to-day activities and ability to work of those suffering from the condition.^70^ An estimated 65 million people worldwide were estimated to be suffering from long COVID in January 2023.^71^ Given the morbidity and economic implications of long COVID (e.g. reduction in productivity), mathematical models and cost-effectiveness analyses should incorporate long COVID in order to best inform policy. Our finding that long COVID was only incorporated into seven studies in this review indicates that it is under-represented in existing modelling literature (within the time period covered by our search). Similarly, the concentration of studies in particular countries and regions (although limited here to high-income countries) signals a need for more modelling studies investigating the impact of vaccination and waning immunity in other areas, which differ in their population structure and healthcare systems. Latest evidence on the duration of immunity and protection against reinfection must be incorporated into future models to ensure reliability of their predictions. Finally, in the event of emergence of a new variant of concern, the results of modelling studies that have explored such a scenario could help to inform quick policy decisions on the most effective disease control strategies to employ.

This is the first review of dynamic transmission, mathematical models of SARS-CoV-2 addressing vaccination and waning immunity to compare model methodologies. We identified gaps in the literature in order to support future model development. Included studies generally scored highly in the quality assessment, increasing the reliability of our study conclusions. A limitation of this review arises from the fact that we did not extract study objectives: that is, that we were not able to compare the model features in the context of the research aims and conclusions. Therefore, this review does not discuss the strengths and weaknesses of each modelling approach specifically in relation to each public health question, nor does it assess the validity and accuracy of model predictions. Our restrictions in terms of eligibility criteria also mean that we cannot comment on the entire range of existing mathematical models, particularly in low- to upper middle-income settings, and any shortage of model types that we have identified here may not exist within the wider literature. A further limitation of this review is that there may be eligible models that were not identified in the database searches and have not been included. We aimed to address this limitation by scanning the reference lists of included studies and searching for articles describing the European COVID-19 Scenario Hub models, to capture any which may have been missed. Incorporating the additional seven studies found through this approach did not change our conclusions.

Other systematic reviews of COVID-19 modelling not focussed on the methodology have also found that most models were deterministic and compartmental, and that outcomes were centred around case numbers, hospital utilisation and deaths, and rarely included QALYs.^12,15,16^ Conversely, stochastic, agent-based models were more common in a systematic review of models in healthcare settings.^13^ To our knowledge, our study is the first to describe the variety of existing SARS-CoV-2 modelling methods and features in such detail. Following the rapid, urgent development of many different transmission models after SARS-CoV-2 first emerged, it is important to reflect upon what types of models are needed in future. This review provides the perspective needed to do so and may be of use to those wishing to assess modelling methodology more closely.

Within the setting of high-income countries, our results highlight certain considerations required for future disease control planning that should have increased representation in mathematical modelling studies. Most importantly, there is a clear need for more long-term models that include long COVID, measures of disease burden that include quality of life, the potential impact of novel variants, or an economic evaluation of control strategies. The findings from this review provide critical information for policymakers and infectious disease modellers who are considering what to prioritise when evaluating or producing a new model.

A broader systematic review of SARS-CoV-2 modelling studies in non high-income settings, and that is not limited to age-structured models of vaccination and waning immunity, would allow further analysis into the range of existing literature; however, this was beyond the scope of the current study. Future modelling studies looking to inform policy decisions should focus on incorporating the elements we have identified above: long COVID, QALYs, novel variants and economic outcomes.

## Declaration of interests

### Contributors

HC, EBP, and EB conceived and designed the study; HC, EBP and ZW supervised the study. EB ran the database searches and de-duplicated the results. EB and SAK conducted the title and abstract screening; EB, SAK, JS and JD conducted the full text screening. All authors extracted data and took part in the quality assessment process. EB interpreted the data and wrote the first draft of the manuscript. All authors reviewed and revised the manuscript and approved the final version.

## Data Availability

All data produced in the present study are available upon reasonable request to the authors

## Acknowledgements

HC, EBP, ZW and EB acknowledge support from the National Institute for Health Research Health Protection Research Unit (NIHR HPRU) in Behavioural Science and Evaluation at the University of Bristol in partnership with Public Health England. HC is supported by a NIHR Career Development Fellowship (CDF-2018-11-ST2-015). SAK is supported in part by grant MR/N0137941/1 for the GW4 BioMed MRC DTP, awarded to the Universities of Bath, Bristol, Cardiff and Exeter from the Medical Research Council (MRC)/UKRI. JS acknowledges funding from the Wellcome Trust [WT 226619/Z/22/Z]. JD is funded in part by grant MR/W006308/1 for the GW4 BIOMED MRC DTP, awarded for the Universities of Bath, Bristol, Cardiff and Exeter from the Medical Research Council (MRC)/UKRI. ZW is supported by a National Institute for Health Research (NIHR) Health Technology Health Assessment grant (NIHR128513). We thank Oliver Watson, Thu Tran, Joshua Szanyi, Matt Keeling, Samuel Alizon, Paula Sanz-Leon, Chloe Bracis, Andrea Cattaneo, Sabina Marchetti, Katie Mintram, Frank Sandmann, Alexander Beams, Bruce Lee, Marie Ferguson Martinez, Seyed Moghadas, Simon Cauchemez and Paolo Bosetti for providing further information for this review.

## Statement of ethical approval

Due to the nature of this study (a systematic review), ethical approval was not required.

## References

1. UKHSA. COVID-19 vaccine surveillance report: 12 January 2023 (week 2) [Internet]. 2023 [cited 2024 Jan 31]. Available from: https://www.gov.uk/government/publications/covid-19-vaccine-weekly-surveillance-reports

2. Andrews N, Tessier E, Stowe J, Gower C, Kirsebom F, Simmons R, et al. Duration of Protection against Mild and Severe Disease by Covid-19 Vaccines. New England Journal of Medicine [Internet]. 2022 Jan 27 [cited 2023 Sep 18];386(4):340–50. Available from: 10.1056/NEJMoa2115481

3. Ferguson N, Ghani A, Cori A, Hogan A, Hinsley W, Volz E. Growth, population distribution and immune escape of the Omicron in England. Imperial College London [Internet]. 2021. Available from: 10.25561/93038

4. Brooks-Pollock E, Danon L, Jombart T, Pellis L. Modelling that shaped the early COVID-19 pandemic response in the UK. Philosophical Transactions of the Royal Society B [Internet]. 2021 Jul 19 [cited 2024 Jan 24];376(1829). Available from: 10.1098/rstb.2021.0001

5. Jentsch PC, Anand M, Bauch CT. Prioritising COVID-19 vaccination in changing social and epidemiological landscapes: a mathematical modelling study. Lancet Infect Dis [Internet]. 2021 Aug 1 [cited 2024 Jan 24];21(8):1097–106. Available from: 10.1016/S1473-3099(21)00057-8

6. Hill EM, Keeling MJ. Comparison between one and two dose SARS-CoV-2 vaccine prioritization for a fixed number of vaccine doses. J R Soc Interface [Internet]. 2021 [cited 2024 Jan 24];18(182). Available from: 10.1098/rsif.2021.0214

7. Moore S, Hill EM, Dyson L, Tildesley MJ, Keeling MJ. Modelling optimal vaccination strategy for SARS-CoV-2 in the UK. PLoS Comput Biol [Internet]. 2021 May 1 [cited 2024 Jan 24];17(5):e1008849. Available from: 10.1371/journal.pcbi.1008849

8. MacIntyre CR, Costantino V, Trent M. Modelling of COVID-19 vaccination strategies and herd immunity, in scenarios of limited and full vaccine supply in NSW, Australia. Vaccine [Internet]. 2022 Apr 14 [cited 2023 Oct 27];40(17):2506–13. Available from: 10.1016/J.VACCINE.2021.04.042

9. Bubar KM, Reinholt K, Kissler SM, Lipsitch M, Cobey S, Grad YH, et al. Model-informed COVID-19 vaccine prioritization strategies by age and serostatus. Science (1979) [Internet]. 2021 Feb 26 [cited 2024 Jan 24];371(6532):916–21. Available from: 10.1126/science.abe6959

10. Lin YF, Duan Q, Zhou Y, Yuan T, Li P, Fitzpatrick T, et al. Spread and impact of COVID-19 in China: A systematic review and synthesis of predictions from transmission-dynamic models. Front Med (Lausanne) [Internet]. 2020 Jun 18 [cited 2023 Oct 27];7:1–11. Available from: 10.3389/fmed.2020.00321

11. Jinxing G, Yongyue W, Yang Z, Feng C, Jinxing G, Yongyue W, et al. Modeling the transmission dynamics of COVID-19 epidemic: a systematic review. The Journal of Biomedical Research, 2020, Vol 34, Issue 6, Pages: 422-430 [Internet]. 2020 Nov 28 [cited 2023 Oct 27];34(6):422–30. Available from: http://www.jbr-pub.org.cn/en/article/doi/10.7555/JBR.34.20200119

12. Xiang Y, Jia Y, Chen L, Guo L, Shu B, Long E. COVID-19 epidemic prediction and the impact of public health interventions: A review of COVID-19 epidemic models. Infect Dis Model [Internet]. 2021 Jan 1 [cited 2023 Oct 27];6:324–42. Available from: 10.1016/j.idm.2021.01.001

13. Smith DRM, Chervet S, Pinettes T, Shirreff G, Jijón S, Oodally A, et al. How have mathematical models contributed to understanding the transmission and control of SARS-CoV-2 in healthcare settings? A systematic search and review. Journal of Hospital Infection [Internet]. 2023 Nov 1 [cited 2023 Oct 27];141:132–41. Available from: 10.1016/j.jhin.2023.07.028

14. Gerlee P, Jöud A, Spreco A, Timpka T. Computational models predicting the early development of the COVID-19 pandemic in Sweden: systematic review, data synthesis, and secondary validation of accuracy. Scientific Reports 2022 12:1 [Internet]. 2022 Aug 2 [cited 2023 Oct 27];12(1):1–10. Available from: https://doi-org.bris.idm.oclc.org/10.1038/s41598-022-16159-6

15. Saadi N, Chi YL, Ghosh S, Eggo RM, McCarthy C V., Quaife M, et al. Models of COVID-19 vaccine prioritisation: a systematic literature search and narrative review. BMC Med [Internet]. 2021 Dec 1 [cited 2023 Oct 27];19(1):1–11. Available from: 10.1186/s12916-021-02190-3

16. Shankar S, Mohakuda SS, Kumar A, Nazneen PS, Yadav AK, Chatterjee K, et al. Systematic review of predictive mathematical models of COVID-19 epidemic. Med J Armed Forces India [Internet]. 2021 Jul 1 [cited 2023 Oct 27];77:S385–92. Available from: 10.1016/j.mjafi.2021.05.005

17. Tracy M, Cerdá M, Keyes KM. Agent-Based Modeling in Public Health: Current Applications and Future Directions. Annu Rev Public Health [Internet]. 2018 Apr 2 [cited 2024 Jan 31];39:77–94. Available from: 10.1146/annurev-publhealth-040617-014317

18. World Bank Country and Lending Groups – World Bank Data Help Desk [Internet]. [cited 2023 Oct 28]. Available from: https://datahelpdesk.worldbank.org/knowledgebase/articles/906519-world-bank-country-and-lending-groups

19. Ouzzani M, Hammady H, Fedorowicz Z, Elmagarmid A. Rayyan-a web and mobile app for systematic reviews. Syst Rev [Internet]. 2016 Dec 5 [cited 2023 Oct 28];5(1):1–10. Available from: 10.1186/s13643-016-0384-4

20. Harris RC, Sumner T, Knight GM, White RG. Systematic review of mathematical models exploring the epidemiological impact of future TB vaccines. Hum Vaccin Immunother [Internet]. 2016 Nov 1 [cited 2023 Oct 28];12(11):2813–32. Available from: 10.1080/21645515.2016.1205769

21. Koslow W, Kühn MJ, Binder S, Klitz M, Abele D, Basermann A, et al. Appropriate relaxation of non-pharmaceutical interventions minimizes the risk of a resurgence in SARS-CoV-2 infections in spite of the Delta variant. PLoS Comput Biol [Internet]. 2022 May 1 [cited 2023 Oct 31];18(5):e1010054. Available from: 10.1371/journal.pcbi.1010054

22. Massonnaud CR, Roux J, Colizza V, Crépey P. Evaluating COVID-19 Booster Vaccination Strategies in a Partially Vaccinated Population: A Modeling Study. Vaccines (Basel) [Internet]. 2022 Mar 1 [cited 2023 Oct 31];10(3):479. Available from: 10.3390/vaccines10030479

23. Rodiah I, Vanella P, Kuhlmann A, Jaeger VK, Harries M, Krause G, et al. Age-specific contribution of contacts to transmission of SARS-CoV-2 in Germany. Eur J Epidemiol [Internet]. 2023 Jan 1 [cited 2023 Oct 31];38(1):39–58. Available from: 10.1007/s10654-022-00938-6

24. Tran TNA, Wikle NB, Albert E, Inam H, Strong E, Brinda K, et al. Optimal SARS-CoV-2 vaccine allocation using real-time attack-rate estimates in Rhode Island and Massachusetts. BMC Med [Internet]. 2021 Dec 1 [cited 2023 Oct 31];19(1):1–14. Available from: 10.1186/s12916-021-02038-w

25. Makhoul M, Chemaitelly H, Ayoub HH, Seedat S, Abu-Raddad LJ. Epidemiological Differences in the Impact of COVID-19 Vaccination in the United States and China. Vaccines 2021, Vol 9, Page 223 [Internet]. 2021 Mar 5 [cited 2023 Oct 31];9(3):223. Available from: 10.3390/vaccines9030223

26. Marziano V, Guzzetta G, Mammone A, Riccardo F, Poletti P, Trentini F, et al. The effect of COVID-19 vaccination in Italy and perspectives for living with the virus. Nature Communications 2021 12:1 [Internet]. 2021 Dec 14 [cited 2023 Oct 31];12(1):1–8. Available from: 10.1038/s41467-021-27532-w

27. Mendes D, Chapman R, Gal P, Atwell J, Nguyen JL, Hamson L, et al. Public health impact of booster vaccination against COVID-19 in the UK during Delta variant dominance in autumn 2021. J Med Econ [Internet]. 2022 Dec 31 [cited 2023 Oct 31];25(1):1039–50. Available from: 10.1080/13696998.2022.2111935

28. Moore S, Hill EM, Dyson L, Tildesley MJ, Keeling MJ. Retrospectively modeling the effects of increased global vaccine sharing on the COVID-19 pandemic. Nature Medicine 2022 28:11 [Internet]. 2022 Oct 27 [cited 2023 Oct 31];28(11):2416–23. Available from: 10.1038/s41591-022-02064-y

29. Carlsson M, Söderberg-Nauclér C. COVID-19 Modeling Outcome versus Reality in Sweden. Viruses [Internet]. 2022 Aug 1 [cited 2023 Oct 31];14(8):1840. Available from: 10.3390/v14081840

30. Sofonea MT, Roquebert B, Foulongne V, Morquin D, Verdurme L, Trombert-Paolantoni S, et al. Analyzing and Modeling the Spread of SARS-CoV-2 Omicron Lineages BA.1 and BA.2, France, September 2021–February 2022 - Volume 28, Number 7—July 2022 - Emerging Infectious Diseases journal - CDC. Emerg Infect Dis [Internet]. 2022 Jul 1 [cited 2023 Oct 31];28(7):1355–65. Available from: 10.3201/eid2807.220033

31. Sandmann FG, Davies NG, Vassall A, Edmunds WJ, Jit M, Sun FY, et al. The potential health and economic value of SARS-CoV-2 vaccination alongside physical distancing in the UK: a transmission model-based future scenario analysis and economic evaluation. Lancet Infect Dis [Internet]. 2021 Jul 1 [cited 2023 Oct 31];21(7):962–74. Available from: 10.1016/S1473-3099(21)00079-7

32. Coudeville L, Jollivet O, Mahé C, Chaves S, Gomez GB. Potential impact of introducing vaccines against COVID-19 under supply and uptake constraints in France: A modelling study. PLoS One [Internet]. 2021 Apr 1 [cited 2023 Oct 31];16(4):e0250797. Available from: 10.1371/journal.pone.0250797

33. Beams AB, Bateman R, Adler FR. Will SARS-CoV-2 Become Just Another Seasonal Coronavirus? Viruses 2021, Vol 13, Page 854 [Internet]. 2021 May 7 [cited 2023 Oct 31];13(5):854. Available from: 10.3390/v13050854

34. Aruffo E, Yuan P, Tan Y, Gatov E, Moyles I, Bélair J, et al. Mathematical modelling of vaccination rollout and NPIs lifting on COVID-19 transmission with VOC: a case study in Toronto, Canada. BMC Public Health [Internet]. 2022 Dec 1 [cited 2023 Oct 31];22(1):1–12. Available from: 10.1186/s12889-022-13597-9

35. Barnard RC, Davies NG, Munday JD, Lowe R, Knight GM, Leclerc QJ, et al. Modelling the medium-term dynamics of SARS-CoV-2 transmission in England in the Omicron era. Nature Communications 2022 13:1 [Internet]. 2022 Aug 19 [cited 2023 Oct 31];13(1):1–15. Available from: 10.1038/s41467-022-32404-y

36. Bartsch SM, O’Shea KJ, Chin KL, Strych U, Ferguson MC, Bottazzi ME, et al. Maintaining face mask use before and after achieving different COVID-19 vaccination coverage levels: a modelling study. Lancet Public Health [Internet]. 2022 Apr 1 [cited 2023 Oct 31];7(4):e356–65. Available from: 10.1016/S2468-2667(22)00040-8

37. Bracis C, Moore M, Swan DA, Matrajt L, Anderson L, Reeves DB, et al. Improving vaccination coverage and offering vaccine to all school-age children allowed uninterrupted in-person schooling in King County, WA: Modeling analysis. Mathematical Biosciences and Engineering 2022 6:5699 [Internet]. 2022 [cited 2023 Oct 31];19(6):5699–716. Available from: 10.3934/mbe.2022266

38. Childs L, Dick DW, Feng Z, Heffernan JM, Li J, Röst G. Modeling waning and boosting of COVID-19 in Canada with vaccination. Epidemics [Internet]. 2022 Jun 1 [cited 2023 Oct 31];39. Available from: 10.1016/j.epidem.2022.100583

39. Chun JY, Jeong H, Kim Y. Identifying susceptibility of children and adolescents to the Omicron variant (B.1.1.529). BMC Med [Internet]. 2022 Dec 1 [cited 2023 Oct 31];20(1):1–9. Available from: 10.1186/s12916-022-02655-z

40. Dick DW, Childs L, Feng Z, Li J, Röst G, Buckeridge DL, et al. Covid-19 seroprevalence in canada modelling waning and boosting covid-19 immunity in canada a canadian immunization research network study. Vaccines (Basel) [Internet]. 2022 Jan 1 [cited 2023 Oct 31];10(1):17. Available from: 10.3390/vaccines10010017

41. Glasser JW, Feng Z, Vo M Van, Jones JN, Clarke KEN. Analysis of serological surveys of antibodies to SARS-CoV-2 in the United States to estimate parameters needed for transmission modeling and to evaluate and improve the accuracy of predictions. J Theor Biol [Internet]. 2023 Jan 7 [cited 2023 Oct 31];556:111296. Available from: 10.1016/j.jtbi.2022.111296

42. Hogan AB, Winskill P, Watson OJ, Walker PGT, Whittaker C, Baguelin M, et al. Within-country age-based prioritisation, global allocation, and public health impact of a vaccine against SARS-CoV-2: A mathematical modelling analysis. Vaccine [Internet]. 2021 May 21 [cited 2023 Oct 31];39(22):2995–3006. Available from: 10.1016/j.vaccine.2021.04.002

43. Keeling MJ, Moore SE. An assessment of the vaccination of school-aged children in England against SARS-CoV-2. BMC Med [Internet]. 2022 Dec 1 [cited 2023 Oct 31];20(1):1–14. Available from: 10.1186/s12916-022-02379-0

44. Kraay ANM, Gallagher ME, Ge Y, Han P, Baker JM, Koelle K, et al. The role of booster vaccination and ongoing viral evolution in seasonal circulation of SARS-CoV-2. J R Soc Interface [Internet]. 2022 Sep 7 [cited 2023 Oct 31];19(194). Available from: 10.1098/rsif.2022.0477

45. Liang JB, Yuan HY, Li KK, Wei WI, Wong SYS, Tang A, et al. Path to normality: Assessing the level of social-distancing measures relaxation against antibody-resistant SARS-CoV-2 variants in a partially-vaccinated population. Comput Struct Biotechnol J [Internet]. 2022 Jan 1 [cited 2023 Oct 31];20:4052–9. Available from: 10.1016/j.csbj.2022.07.048

46. Liu Y, Sandmann FG, Barnard RC, Pearson CAB, Pastore R, Pebody R, et al. Optimising health and economic impacts of COVID-19 vaccine prioritisation strategies in the WHO European Region: a mathematical modelling study. The Lancet Regional Health - Europe [Internet]. 2022 Jan 1 [cited 2023 Oct 31];12:100267. Available from: 10.1016/j.lanepe.2021.100267

47. Yang W, Shaman J. Development of a model-inference system for estimating epidemiological characteristics of SARS-CoV-2 variants of concern. Nature Communications 2021 12:1 [Internet]. 2021 Sep 22 [cited 2023 Oct 31];12(1):1–9. Available from: 10.1038/s41467-021-25913-9

48. Song F, Bachmann MO. Vaccination against COVID-19 and society’s return to normality in England: a modelling study of impacts of different types of naturally acquired and vaccine-induced immunity. BMJ Open [Internet]. 2021 Nov 1 [cited 2023 Oct 31];11(11):e053507. Available from: 10.1136/bmjopen-2021-053507

49. Caetano C, Morgado ML, Patrício P, Leite A, Machado A, Torres A, et al. Measuring the impact of COVID-19 vaccination and immunity waning: A modelling study for Portugal. Vaccine [Internet]. 2022 Nov 22 [cited 2023 Oct 31];40(49):7115–21. Available from: 10.1016/j.vaccine.2022.10.007

50. Watson OJ, Barnsley G, Toor J, Hogan AB, Winskill P, Ghani AC. Global impact of the first year of COVID-19 vaccination: a mathematical modelling study. Lancet Infect Dis [Internet]. 2022 Sep 1 [cited 2023 Oct 31];22(9):1293–302. Available from: 10.1016/S1473-3099(22)00320-6

51. Roy J, Heath SM, Wang S, Ramkrishna D. Modeling COVID-19 transmission between age groups in the United States considering virus mutations, vaccinations, and reinfection. Scientific Reports 2022 12:1 [Internet]. 2022 Nov 22 [cited 2023 Oct 31];12(1):1–16. Available from: 10.1038/s41598-022-21559-9

52. Sonabend R, Whittles LK, Imai N, Perez-Guzman PN, Knock ES, Rawson T, et al. Non-pharmaceutical interventions, vaccination, and the SARS-CoV-2 delta variant in England: a mathematical modelling study. The Lancet [Internet]. 2021 Nov 13 [cited 2023 Oct 31];398(10313):1825–35. Available from: 10.1016/S0140-6736(21)02276-5

53. Barmpounakis P, Demiris N, Kontoyiannis I, Pavlakis GN, Sypsa V. Evaluating the effects of second-dose vaccine-delay policies in European countries: A simulation study based on data from Greece. PLoS One [Internet]. 2022 Apr 1 [cited 2023 Oct 31];17(4):e0263977. Available from: 10.1371/journal.pone.0263977

54. Ainslie KEC, Backer JA, de Boer PT, van Hoek AJ, Klinkenberg D, Altes HK, et al. A scenario modelling analysis to anticipate the impact of COVID-19 vaccination in adolescents and children on disease outcomes in the Netherlands, summer 2021. Eurosurveillance [Internet]. 2022 Nov 3 [cited 2023 Oct 31];27(44):2101090. Available from: 10.2807/1560-7917.ES.2022.27.44.2101090

55. Sanz-Leon P, Hamilton LHW, Raison SJ, Pan AJX, Stevenson NJ, Stuart RM, et al. Modelling herd immunity requirements in Queensland: impact of vaccination effectiveness, hesitancy and variants of SARS-CoV-2. Philosophical Transactions of the Royal Society A [Internet]. 2022 Oct 3 [cited 2023 Oct 31];380(2233). Available from: 10.1098/rsta.2021.0311

56. Truszkowska A, Zino L, Butail S, Caroppo E, Jiang ZP, Rizzo A, et al. Predicting the Effects of Waning Vaccine Immunity Against COVID-19 through High-Resolution Agent-Based Modeling. Adv Theory Simul [Internet]. 2022 Jun 1 [cited 2023 Oct 31];5(6):2100521. Available from: 10.1002/adts.202100521

57. Szanyi J, Wilson T, Howe S, Zeng J, Andrabi H, Rossiter S, et al. Epidemiologic and economic modelling of optimal COVID-19 policy: public health and social measures, masks and vaccines in Victoria, Australia. Lancet Reg Health West Pac [Internet]. 2023 Mar 1 [cited 2023 Oct 31];32:100675. Available from: 10.1016/j.lanwpc.2022.100675

58. Singh DE, Olmedo Luceron C, Limia Sanchez A, Guzman Merino M, Duran Gonzalez C, Delgado-Sanz C, et al. Evaluation of vaccination strategies for the metropolitan area of Madrid via agent-based simulation. BMJ Open [Internet]. 2022 Dec 1 [cited 2023 Oct 31];12(12):e065937. Available from: 10.1136/bmjopen-2022-065937

59. Cattaneo A, Vitali A, Mazzoleni M, Previdi F. An agent-based model to assess large-scale COVID-19 vaccination campaigns for the Italian territory: The case study of Lombardy region. Comput Methods Programs Biomed [Internet]. 2022 Sep 1 [cited 2023 Oct 31];224:107029. Available from: 10.1016/j.cmpb.2022.107029

60. Mintram K, Anagnostou A, Anokye N, Okine E, Groen D, Saha A, et al. CALMS: Modelling the long-term health and economic impact of Covid-19 using agent-based simulation. PLoS One [Internet]. 2022 Aug 1 [cited 2023 Oct 31];17(8):e0272664. Available from: 10.1371/journal.pone.0272664

61. Yang W, Greene SK, Peterson ER, Li W, Mathes R, Graf L, et al. Epidemiological characteristics of the B.1.526 SARS-CoV-2 variant. Sci Adv [Internet]. 2022 Jan 1 [cited 2024 Jan 7];8(4). Available from: 10.1126/sciadv.abm0300

62. Moghadas SM, Vilches TN, Zhang K, Nourbakhsh S, Sah P, Fitzpatrick MC, et al. Evaluation of COVID-19 vaccination strategies with a delayed second dose. PLoS Biol [Internet]. 2021 Apr 1 [cited 2024 Jan 7];19(4):e3001211. Available from: 10.1371/journal.pbio.3001211

63. Panovska-Griffiths J, Swallow B, Hinch R, Cohen J, Rosenfeld K, Stuart RM, et al. Statistical and agent-based modelling of the transmissibility of different SARS-CoV-2 variants in England and impact of different interventions. Philosophical Transactions of the Royal Society A [Internet]. 2022 Oct 3 [cited 2024 Jan 7];380(2233). Available from: 10.1098/rsta.2021.0315

64. Li R, Bjørnstad ON, Stenseth NC. Switching vaccination among target groups to achieve improved long-lasting benefits. R Soc Open Sci [Internet]. 2021 Jun 16 [cited 2024 Jan 7];8(6). Available from: 10.1098/rsos.210292

65. Gavish N, Yaari R, Huppert A, Katriel G. Population-level implications of the Israeli booster campaign to curtail COVID-19 resurgence. Sci Transl Med [Internet]. 2022 Jun 1 [cited 2024 Jan 7];14(647):9836. Available from: 10.1126/scitranslmed.abn9836

66. Bosetti P, Kiem CT, Andronico A, Paireau J, Levy-Bruhl D, Alter L, et al. Impact of booster vaccination on the control of COVID-19 Delta wave in the context of waning immunity: application to France in the winter 2021/22. Eurosurveillance [Internet]. 2022 Jan 6 [cited 2024 Jan 7];27(1):2101125. Available from: 10.2807/1560-7917.ES.2022.27.1.2101125

67. Shattock AJ, Le Rutte EA, Dünner RP, Sen S, Kelly SL, Chitnis N, et al. Impact of vaccination and non-pharmaceutical interventions on SARS-CoV-2 dynamics in Switzerland. Epidemics [Internet]. 2022 Mar 1 [cited 2023 Oct 31];38:100535. Available from: 10.1016/j.epidem.2021.100535

68. Hernandez-Suarez C, Murillo-Zamora E. Waning immunity to SARS-CoV-2 following vaccination or infection. Front Med (Lausanne) [Internet]. 2022 Oct 13 [cited 2023 Nov 3];9:972083. Available from: 10.3389/fmed.2022.972083

69. Stein C, Nassereldine H, Sorensen RJD, Amlag JO, Bisignano C, Byrne S, et al. Past SARS-CoV-2 infection protection against re-infection: a systematic review and meta-analysis. The Lancet [Internet]. 2023 Mar 11 [cited 2024 Jan 12];401(10379):833–42. Available from: 10.1016/S0140-6736(22)02465-5

70. Gualano MR, Rossi MF, Borrelli I, Santoro PE, Amantea C, Daniele A, et al. Returning to work and the impact of post COVID-19 condition: A systematic review. Work [Internet]. 2022 Jan 1 [cited 2023 Nov 3];73(2):405–13. Available from: 10.3233/WOR-220103

71. Davis HE, McCorkell L, Vogel JM, Topol EJ. Long COVID: major findings, mechanisms and recommendations. Nature Reviews Microbiology 2023 21:3 [Internet]. 2023 Jan 13 [cited 2024 Jan 16];21(3):133–46. Available from: 10.1038/s41579-022-00846-2

