## Supplementary file 1 for "Mathematical models of COVID-19 vaccination in high-income countries: A systematic review"

Supplementary File 1 – Mathematical models of COVID-19 vaccination: a systematic review

**Methods**

*Assumptions and details of data extraction*

If a study made no mention of an economic evaluation or of costs in the model then it was assumed this was not conducted. Similarly, if immune escape or a delay between vaccination and development of immunity were not described then these were assumed to be not included, and if the paper did not state whether the model was deterministic or stochastic but there was no description of stochastic methods then it was assumed to be deterministic. For the time horizon of the study, we extracted the maximum time period over which model analysis was carried out (excluding the fitting period for predictive studies). When reporting the contact matrix structure, we consider only the contact matrix used in the main model and not in sensitivity analyses. We defined the number of doses modelled as the maximum number of doses an individual could have received by the end of the simulated period. For the delay between vaccination and development of immunity, we reported the time delay following the second dose in each model (including the time to peak protection when there was a gradual increase in immunity over time since vaccination).

**Search strategy**

We carried out a search of PubMed, Scopus and Embase on the 6^th^ February 2023. The search was restricted to publications from the year 2019 until the date it was conducted, and articles were limited to peer-reviewed papers published in English. Reference lists of included papers were also checked for other relevant studies.

We used the following search terms for each database:

PubMed

*(("COVID-19"[Mesh] OR "SARS-CoV-2"[Mesh] OR (covid 19[Title]) OR (sars cov 2[Title]) OR (coronavirus[Title]))) AND (("COVID-19 Vaccines"[Mesh] OR "Vaccination"[Mesh] OR "Immunization"[Mesh] OR (vaccin*[Title]) OR (immunisation*[Title]) OR (immunization*[Title]))) AND ("Epidemiological Models"[Mesh:NoExp] OR "Models, Theoretical"[Mesh:NoExp] OR (model*[Title]) OR (simulat*[Title])) AND ((english[Filter]) AND (2019:2023[pdat])) AND (humans[Filter]) NOT ("animal model*"[Title] OR "molecular model*"[Title] OR "health belief model*"[Title] OR mouse[Title] OR hamster*[Title]) NOT (animal[Filter]) NOT (preprint[Filter] OR review[Filter])*

Scopus

*( ( TITLE ( "covid 19" ) OR TITLE ( "SARS COV 2" ) OR TITLE ( coronavirus ) ) AND ( TITLE ( vaccin* ) OR TITLE ( immunisation* ) OR TITLE ( immunization* ) ) AND ( TITLE ( model* ) OR TITLE ( simulat* ) ) ) AND NOT ( TITLE ( "animal model*" ) OR TITLE ( "molecular model*" ) OR TITLE ( "health belief model*" ) OR TITLE ( mouse ) OR TITLE ( hamster* ) ) AND ( LIMIT-TO ( PUBSTAGE , "final" ) ) AND ( LIMIT-TO ( DOCTYPE , "ar" ) ) AND ( LIMIT-TO ( PUBYEAR , 2023 ) OR LIMIT-TO ( PUBYEAR , 2022 ) OR LIMIT-TO ( PUBYEAR , 2021 ) OR LIMIT-TO ( PUBYEAR , 2020 ) OR LIMIT-TO ( PUBYEAR , 2019 ) ) AND ( LIMIT-TO ( LANGUAGE , "English" ) )*

Embase

*((exp coronavirus disease 2019/ or exp Severe acute respiratory syndrome coronavirus 2/ or (covid 19 or sars cov 2 or coronavirus).m_titl.) and (exp SARS-CoV-2 vaccine/ or vaccination/ or immunization/ or (vaccin* or immunisation* or immunization*).m_titl.) and (mathematical model/ or theoretical model/ or exp epidemiological model/ or exp compartment model/ or (model* or simulat*).m_titl.)) not (animal model* or molecular model* or health belief model* or mouse or hamster*).m_titl.*

*limit to (human and english language and yr="2019 -Current" and article)*

**Quality assessment tool**

| **Title:** |
| --- |
| **Authors:** |
| **DOI:** |

|  | **Criterion** | | **Considerations** | **Score considerations (0, poor to 2, good)** | | **Score** | **Total score** |
| --- | --- | --- | --- | --- | --- | --- | --- |
| 1 | **Are the aims and objectives clear?** | | Are the research questions and modelling objectives clearly defined? | 0 Not stated  1 Stated but vague  2 Stated and focussed | |  | Definitions (max 8 points): |
| 2 | **Is the setting and population clearly defined?** | | Does the paper clearly state the setting (e.g. geographical location, care facility)? | 0 Not stated  1 Stated but vague or details missing  2 Stated and focussed | |  |  |
|  |  |  | In health economics models, has the perspective been stated? |  |  |  |  |
|  |  |  | Does the paper clearly state the modelled population? (e.g. patient or population group characteristics) |  |  |  |  |
|  |  |  | Have sub-populations necessary for the research question and setting been modelled? |  |  |  |  |
| 3 | **Are the intervention and comparators adequately defined?** | | Does the paper clearly state the population(s) targeted for vaccination? | 0 Not stated or very unclear  1 Stated but details missing  2 Stated and all necessary details stated | |  |  |
|  |  |  | Does the paper clearly define the vaccine characteristics (e.g. vaccine efficacy, duration of protection, number of doses, waning, timing, delay between vaccination and immunity, type of vaccine used, assumed vaccine uptake levels)? |  |  |  |  |
|  |  |  | If there is a comparator (no vaccine, baseline or alternative intervention scenario), is it clearly defined? |  |  |  |  |
| 4 | **Are the outcome measures defined and answer the research question?** | | Does the paper clearly define the outcomes of interest? | 0 Not stated, very unclear or not suited to research question  1 Stated but details missing or not directly aligned with research question  2 Stated, all necessary details stated, and aligned with research question | |  |  |
|  |  |  | Do the outcomes correspond to the research question? |  |  |  |  |
| 5 | **Are the model structure and time horizon clearly described and appropriate for the research question?** | | Is the model structure clearly reported and appropriate for the research question? | 0 Not appropriate model structure, or poor/no description of model  1 Incomplete description, and/or appropriate in part for research question  2 Complete and reproducible, appropriate structure and time horizon | |  | Model methods (max 4 points): |
|  |  |  | Does the model reflect current knowledge of disease natural history (e.g. duration of immunity, immune escape)? |  |  |  |  |
|  |  |  | Is the time horizon and time step of the model clearly stated and appropriate to the research question (i.e. is it long enough to capture health effects)? |  |  |  |  |
| 6 | **Are the modelling methods appropriate for the research question and adequately described?** | | Were the modelling methods clearly described, and suited to the research question? | 0 Not appropriate model structure, or poor/no description of methods  1 Incomplete description, and/or appropriate in part for research question  2 Complete and reproducible, appropriate method | |  |  |
| 7 | **Are the parameters, ranges and data sources specified?** | | Are all parameters and their ranges reported? | 0 Poorly reported  1 Some information missing  2 Complete reporting of parameters, ranges and data sources | |  | Model inputs (max 6 points): |
|  |  |  | Are the data sources for parameters reported? |  |  |  |  |
| 8 | **Are any assumptions explicit and justified?** | | Are all assumptions explicit and justified? | 0 Not reported  1 Explicit  2 Explicit and justified | |  |  |
| 9 | **Is the quality of data considered and is uncertainty explored through uncertainty and/or sensitivity analyses?** | | Are data limitations discussed? Are any of the sources known to the reviewer to be inappropriate (e.g. do not match the parameter, are outdated, or known to be poor quality)? | 0 No sources or uncertainty  1 Partially addressed, and/or some data inappropriate  2 Fully addressed | |  |  |
|  |  |  | Is uncertainty in model structure, parameters and/or assumptions explored through uncertainty and/or sensitivity analyses? |  |  |  |  |
| 10 | **Is the method of fitting described and suitable?** | | Is the method of fitting/calibrating the model clearly described? | 0 Not done, unsuitable method or poor/no description  1 Incomplete description or method not optimal  2 Complete description and suitable methods | |  | Fitting/ validation (max 4 points): |
|  |  |  | Is the method of model fitting/calibration suitable? |  |  |  |  |
| 11 | **Has the model been validated?** | | Has an assessment of validity of the results been made by comparing across one or more different model structures, or against a validation data set? | 0 Not considered  1 States criteria for validation  2 Validation undertaken | |  |  |
| 12 | **Have the results been clearly and completely presented, with a range of uncertainty?** | | Have the outcome values and their uncertainty ranges for each intervention/scenario been reported? | 0 Not reported, very unclear or not suited to research question  1 Stated, but ranges or planned sensitivity analyses missing and/or not directly aligned with research question  2 Values and ranges and planned sensitivity analyses reported and aligned with research question. | |  | Results (max 4 points): |
|  |  |  | Do the results match the objectives? |  |  |  |  |
|  |  |  | Are sensitivity analyses clearly reported? |  |  |  |  |
| 13 | **Are the results appropriately interpreted and discussed in context?** | | Does the discussion reflect a fair and balanced interpretation of the results? | 0 No/poor discussion  1 Some discussion but key points, limitations or context missed  2 Full discussion of key points in context, generalisability considered, limitations discussed | |  |  |
|  |  |  | Are the results of the study discussed in context and is generalisability considered? |  |  |  |  |
|  |  |  | Are possible biases and limitations discussed? |  |  |  |  |
| 14 | **Are the funding source and conflicts of interest reported?** | | Is the funding and the role of the funder clearly stated? | 0 No statement of funding or conflicts  1 Funding or conflicts reported  2 Funding and conflict statement | |  | Conflicts (max 2 points): |
|  |  |  | Is there a conflict of interest statement? |  |  |  |  |
| **Overall Score (max 28 points):** | |  |  |  |  | | |
| Very high | | >22 |  |  |  | | |
| High | | 19-22 |  |  |  | | |
| Medium | | .14-18 |  |  |  | | |
| Low | | <14 |  |  |  | | |

This tool was adapted from the tool developed by Harris et al. as part of their systematic review of models of TB vaccines.

Rebecca C. Harris, Tom Sumner, Gwenan M. Knight & Richard G. White (2016) Systematic review of mathematical models exploring the epidemiological impact of future TB vaccines, Human Vaccines & Immunotherapeutics, 12:11, 2813-2832, DOI: 10.1080/21645515.2016.1205769
